# The Immediate Effect of COVID-19 Policies on Social Distancing Behavior in the United States

**DOI:** 10.1101/2020.04.07.20057356

**Authors:** Rahi Abouk, Babak Heydari

## Abstract

Anecdotal evidence points to the effectiveness of COVID-19 social distancing policies, however, their effectiveness vis-a-vis what is driven by public awareness and voluntary actions have not been studied. Policy variations across US states create a natural experiment to study the causal impact of each policy. Using a difference-in-differences methodology, location-based mobility, and daily state-level data on COVID-19 tests and confirmed cases, we rank policies based on their effectiveness. We show that statewide stay-at-home orders had the strongest causal impact on reducing social interactions. In contrast, most of the expected impact of more lenient policies were already reaped from non-policy mechanisms. Moreover, stay-at-home policy results in a steady decline in confirmed cases, starting from ten days after implementation and reaching a 37% decrease after fifteen days, consistent with the testing practices and incubation period of the disease.

## Introduction

In the absence of antiviral drugs and vaccines to mitigate the current COVID-19 pandemic, social distancing has been the major mechanism adopted by various impacted countries (Anderson et al., 2020; Lipsitch et al., 2020). These attempts are made, largely, to keep the peak infection level below the resource capacity of healthcare systems and buy time for possible drug and vaccine development.

Reduction in the social contact rate during pandemic outbreaks is driven by a combination of 1) awareness-driven voluntary actions by individuals and businesses and 2) an array of non-pharmaceutical interventions (NPIs) implemented at the national, state, or local level. Research on the 1918 influenza pandemic in different cities in the US points to the role of both these mechanisms in lowering the mortality rate (N. M. Ferguson et al., 2006).

There is strong evidence that social distancing has played a significant role in containing the first wave of the COVID-19 outbreak in China (Chen et al., 2020; Kraemer et al., 2020), and the latest evidence likewise indicates their effectiveness in a number of European countries (Flaxman et al., 2020). However, the relative impact of social awareness versus policy interventions is yet to be determined for the existing outbreak. In addition, several complementary policies were adopted to increase social distancing, some of which might have had unintended consequences. Therefore, identifying the most effective policies could help policymakers respond efficiently to the outbreak. Decoupling these factors is often challenging since, in most countries, the timing and strength of NPIs are highly correlated with public awareness. Moreover, it is crucial to determine which interventions have a significant impact on lowering the contact rate beyond what can be achieved via awareness mechanisms. Overall, the evaluation of these policies could provide valuable lessons, especially for the states that have not yet adopted NPIs. While most countries follow a central policy scheme in the current pandemic, the federal nature of the United States creates a natural experiment setting suitable for addressing these questions. As of this writing, the US federal government has left NPI decisions to individual states, creating a high level of variation in the type and timing of such policies (Adolph et al., 2020) (Figure 1). While there is strong evidence for reduced social contact in the US, not all of these reductions can be attributed to NPIs: mobility data show that people in most states had already started to reduce the time they spend outside their homes before any NPI was implemented (Figure 2). In fact, for some states such as Idaho, Missouri, Wyoming, and the District of Columbia, people’s presence at home had already increased close to a level of saturation before any social distancing policy went into effect. These trends suggest that attributing current reductions in social interaction to policy measures can be misleading, and they further underscore the need for a formal study to disentangle the direct impact of NPIs from other factors such as awareness and spillover effects.

**Figure 1:**
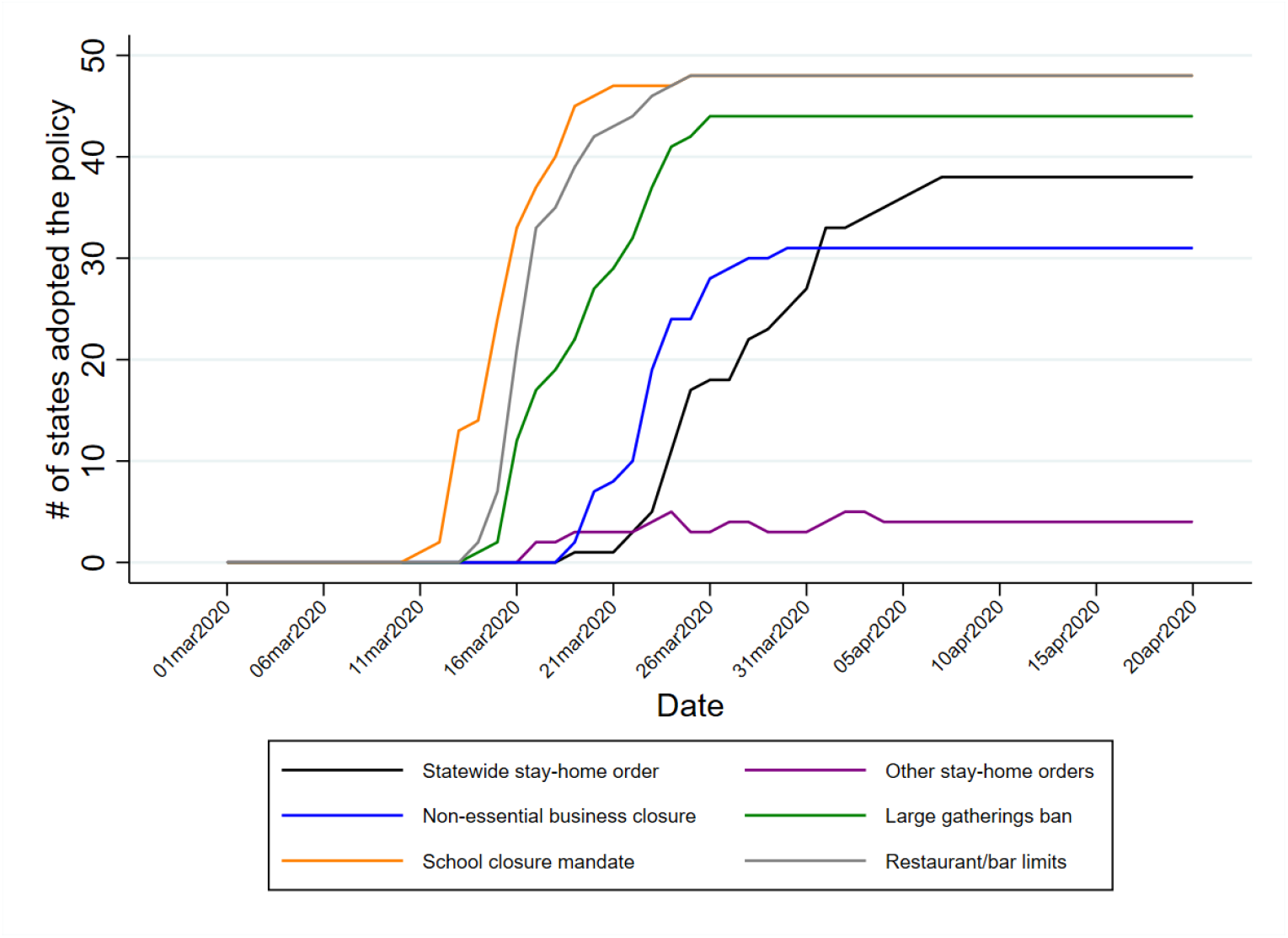
United States COVID-19 policy adoption timeline for six common social distancing policies until March 30 2020.

**Figure 2:**
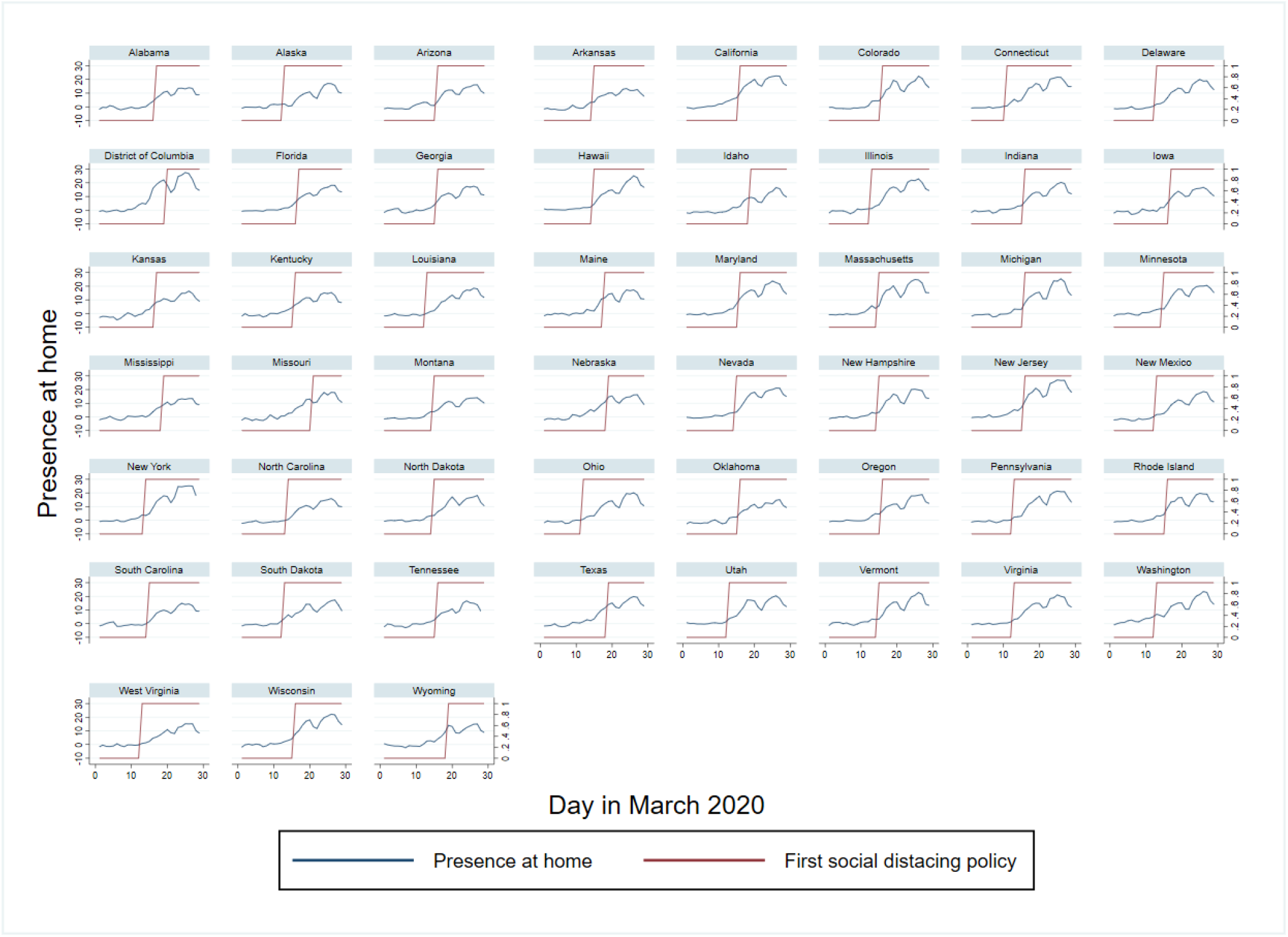
Trends in presence at home and the start date of the first social distancing policy implemented in each state.

This study utilizes the daily state-level variations in the adoption of six different intervention policies–statewide stay-at-home orders, more limited stay-at-home orders, non-essential business closures, large gathering bans, school closure mandates, and restaurant and bar limits–to investigate their causal effects on different indicators of social distancing and on daily positive cases. We first use Google-released daily human mobility indicators for different categories of places, such as residential areas, grocery stores and pharmacies, parks, retail stores and recreation sites, transit stations, and workplaces. The studied intervention policies were each employed in a subgroup of the US states and at different times during March 2020, making them suitable for quasi-experimental methods. We employ a linear regression model and a difference-in-differences approach, which, interestingly, was first developed in a simple form by John Snow in 1849 (Snow, 1855) to study the cause of the Cholera outbreak in London, and it resulted in policy adoptions that effectively ended the outbreak.

Our results give a clear picture of the causal impact and effectiveness of different NPIs on location-specific human mobility in the early stage of the pandemic. At this stage, where changes in behavior are driven by a combination of policy forces and voluntary actions, our results help us rank NPIs based on the strength and significance of their impact, beyond what had already been achieved through voluntary actions. Specifically, our results show that the tendency to remain at home is driven strongly by statewide stay-at-home orders and moderately by non-essential business closures and policies related to restaurant and bar limits. Other policies such as school closure mandates, large gathering bans, and more limited stay-at-home orders do not show any significant impact on keeping people at home. Furthermore, statewide stay-at-home orders have a strong and significant impact on reducing mobility in all outside-home place categories. The effectiveness of non-essential business closures and restaurant/bar limits are also significant on most (but not all) place categories. Limited stay-at-home orders and large gathering bans do not show any impact on increasing presence at home or reducing mobility in other place categories. If anything, in the absence of other policies, large gathering bans result in a moderate increase in mobility at transit stations. School closure mandates give us a mixed picture, with no significant impact on presence at home and a moderate, but significant, impact on mobility in retail locations and transit stations.

Our event study analysis makes a stronger case for the causal nature of these findings. These results show that at the early stages of the pandemic, much of the expected impact of some more lenient NPIs is already achieved by other factors such as voluntary actions and awareness-driven mechanisms. Thus, in order to achieve social distancing beyond what is gained through those mechanisms, states need to adopt strong interventions such as statewide stay-at-home orders.

We then turn to test the causal impact of social distancing policies on the dynamic of the disease based on publicly reported positive test results (confirmed cases). Our approach is similar to what we did for mobility analysis, but here we use a Poisson regression model in our analysis. Our results show that the impact of the statewide stay-at-home order, the strongest NPI based on our findings using community mobility data,start showing reduction ten days after the implementation of the policy and reaches a statistically significant 37% decrease after fifteen days. In contrast, the lenient policies (other stay-at-home orders and large gathering bans) do not result in any statistically significant drop in the growth of the disease.

## Data and Method

### Mobility Trend Data

We use Google-released aggregated, anonymized daily location data on movement trends over time by state, across different categories of places from March 1, 2020 to March 29, 2020 (“COVID-19 Community Mobility Reports”, 2020). The data were gathered by Google from users who have enabled the Location History setting on their accounts and were the same data used by Google Maps to track human traffic at various restaurants and other locations. The data include mobility trends for six location categories including retail and recreation, grocery stores and pharmacies, parks, transit stations, workplaces, and residences. These data were publicly released on April 2, 2020, in the form of charts that plot country-specific mobility trends and the percentage of changes with respect to a baseline for the period of Feb 16 to March 29. The US data also include mobility trends at the level of states and counties. Given that most intervention policies were implemented state-wide, we base our analysis on the state-level data. Each movement trend includes multiple data points per day. We aggregated these points to get a single mobility index per day for each trend chart. We should note that these data do not include people without smartphones, people not carrying their phones to places, etc. However, it is unlikely that the COVID-19 policies affect such changes in recorded behavior. Overall, in most cases we had data on movements for 50 states and the District of Columbia for 29 days, providing us with 1479 observations. However, for some measures, we were not able to restore two observations for a couple of states.

The success of social distancing policies can be evaluated on two fronts. A primary goal of these policies is to decrease the time people spend outside of their homes. In this study, we use the time people spend at residential locations (referred to here as *presence at home* as a proxy to measure the success level of this goal^1^. Besides encouraging people to stay at home, social distancing policies also seek to keep people away from crowded locations and large gatherings, even when they go out. We use the impact of policies on the changes in the remaining five location categories (i.e. retail, transit stations, parks, and workplaces). Given that these categories do not cover all possible gathering places (e.g. places of worship), we base our findings mainly on the impact of policies on presence at home, but we discuss all other categories, acknowledging this limitation.

### State Policy Data

We collected all COVID-19 related policies, their issue dates, and their effective dates for all 50 states and the District of Columbia, going back to the report of the first positive case in the United States. Since there are some discrepancies in policy start dates among datasets available in third-party sources, we used the original documents issued by the state governments, collected by the Kaiser Family Foundation (“State Data and Policy Actions”, 2020), to determine the type and date of each state policy. We considered the effective date as the first day in which the policy in question was in full effect. We acknowledge that this decision creates potential biases since some states had policies that went into effect immediately, resulting in fractions of a day of policy that are missed in our data. This, however, does not substantially change the magnitude and significance of our results.

We performed this study on those policies that aim at social distancing, which we divide into six categories: statewide stay-at-home orders, other stay-at-home orders, non-essential business closures, large gathering bans, school closure mandates, and restaurant and bar limits. *Other stay-at-home orders* incorporates stay-at-home orders for the senior population as well as those targeting specific cities or counties within a given state. Besides these policies, some states have implemented other COVID-19 policies such as cost-sharing waivers for testing or treatment or mandatory quarantine for travelers,which we do not consider in this study. Figure 1 summarizes the policy adoption timeline for each policy by showing the number of states that had each policy in effect on any given day since the beginning of March 2020, suggesting a wide heterogeneity in both the type and the adoption date of each policy during this period.

### Temperature Data

To control for the impact of temperature variation on human mobility, we constructed daily temperature for each state by web-scraping daily temperature data for the top 5 biggest cities in each state from *Weather Underground*, a commercial weather service that provides real-time weather information. We calculated an aggregate the daily temperature for each state by taking the average of daily temperatures of the top 5 cities, weighted by their populations. Table 1 shows the summary statistics for these three categories of data.

**Table 1:** Summary statistics for the community mobility data, March 1, 2020 - March 29, 2020

| Variables | (1)<br>N | (2)<br>Mean | (3)<br>S.D. |
| --- | --- | --- | --- |
| *Mobility data* |  |  |  |
| Presence at home | 1,477 | 6.759 | 7.672 |
| Grocery & pharmacy | 1,478 | 3.180 | 13.590 |
| Parks | 1,477 | 17.340 | 29.350 |
| Retail & recreation | 1,479 | -13.960 | 22.710 |
| Transit stations | 1,479 | -15.070 | 22.260 |
| Workplaces | 1,478 | -16.500 | 18.500 |
| *COVID-19 policies* |  |  |  |
| Statewide stay-at-home order | 1,479 | 0.081 | 0.273 |
| Other stay-at-home orders | 1,479 | 0.026 | 0.160 |
| Non-essential business closure | 1,479 | 0.143 | 0.350 |
| Large gatherings ban | 1,479 | 0.309 | 0.462 |
| School closure mandate | 1,479 | 0.462 | 0.499 |
| Restaurant/bar limits | 1,479 | 0.405 | 0.491 |
| Mean daily temperature (°F) | 1,479 | 48.610 | 12.790 |

Notes: Mobility data are in percentage changes relative to the baseline.

### Positive Cases and Test Data

For the daily state-level number of tests and positive cases (confirmed cases), we employed data from the COVID Tracking Project website (covidtracking.com) from March 9, 2020, to April 20, 2020. We had data on all 50 states and the District of Columbia for 43 days, providing 2193 observations. The primary source for these data is state public health officials. We were aware of minor delays in reporting the number of tests and confirmed cases by states. However, this is an unprecedented and urgent public health issue, and we are only beginning to learn about various aspects of this phenomenon. Therefore, studies such as ours, which can demonstrate the effect of COVID-19 policies, are incredibly valuable for helping policymakers in evaluating alternative scenarios. Table 4 presents the summary statistics for the total COVID-19 tests performed and the number of confirmed cases. On average, between March 9 and April 20, 2020, 26,600 tests were done in each state on a daily basis out of which 351 cased were reported positive.

**Table 4:** Summary statistics for positive test results, March 9, 2020 - April 20, 2020.

| Variables | (1)<br>N | (2)<br>Mean | (3)<br>S.D. |
| --- | --- | --- | --- |
| *Test data* |  |  |  |
| Daily total test results | 2,193 | 26,594 | 55,165 |
| Daily positive tests | 2,193 | 351.3 | 1,045 |
| *COVID-19 policies* |  |  |  |
| Statewide stay-home order | 2,193 | 0.416 | 0.493 |
| Other stay-home orders | 2,193 | 0.0593 | 0.236 |
| Non-essential business closure | 2,193 | 0.407 | 0.491 |
| Large gatherings ban | 2,193 | 0.650 | 0.477 |
| School closure mandate | 2,193 | 0.793 | 0.405 |
| Restaurant/bar limits | 2,193 | 0.755 | 0.430 |
| Mean daily temperature 15 days ago (°F) | 2,193 | 47.42 | 12.87 |

### Difference-in-differences Estimation

Using a linear regression model and a difference-in-differences methodology we evaluate the effect of the COVID-19 policies. In particular, we compare the daily changes in visits from various locations in states that have adopted various COVID 19-related policies with those that have not done so, before and after these policies take effect. For each policy we define a binary variable, set to one if a given state adopts that policy after a certain day during the sample period, and otherwise zero. Note that the validity of this approach assumes parallel trends in changes in visits absent the policies, an assumption which we empirically test using an event study approach. To study the effect of COVID-19 policies, we estimate the following regression equation:

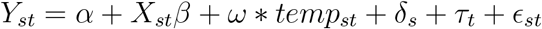

where Y is the changes in visiting various places such as home, grocery and pharmacy, park, retail and recreation, transit station, and workplace. X is the matrix for COVID-19 policies introduced before. *temp* represents state-level mean daily temperature. *δ* and *τ* are sets of state and day-of-the-month fixed effects, respectively. Since changes in visits within the same state are serially correlated over time, we cluster standard errors at the state level (Bertrand et al., 2004). For the event study, we estimate the following regression equation:

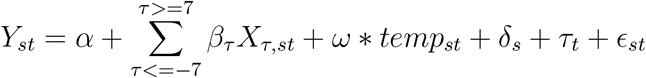

In the event study specification, we replace each policy indicator variable (one at a time) with 26 binary variables, which estimate the effect of that particular policy seven or more days, six to two days (five binary variables), and one day before implementation, as well as one day, two to 14 days (14 binary variables), and 15 days or more after the implementation of the policy. These variables are all zero for states without those policies. We normalize the coefficient for the day before the implementation to zero. Note that the coefficient corresponding to *τ* = *−*1 is normalized to zero and event study for each policy is conducted while including binary variables on other policies in the regression model.

We expect to observe the effect of policies on confirmed cases with delay. Therefore, a simple difference-in-differences estimate is not a suitable option. For this reason, when estimating the effect of COVID-19 policies on the number of confirmed cases, we use the following event study Poisson regression model:

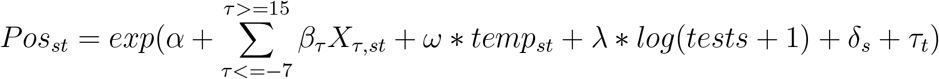

where *Pos*_*st*_ is the state-level daily number of confirmed cases. Given that confirmed cases in each states heavily depends on the number of COVID-19 tests conducted, we also control for this variable in our model. We use a log-transformed version of this variable, which enables us to interpret the estimated coefficient as elasticity. For some days in the sample period, no testings were done in some states. Since log of zero is undefined, we add one. Since the COVID-19 database includes more recent observations, we are able to track the effect of the policies for a longer post-treatment period. In this case, we include 15 lag variables.

## Results

### Impact of Policies on Human Mobility

Table 2 reports the results for the effect of policies introduced above on changes in daily visits from various places. Our main focus is on *presence at home*, for which we report the results in Column (1). Results indicate that statewide stay-at-home orders significantly increase the measure associated with *presence at home* by about six fold (relative to states without such a policy), while more limited stay-at-home orders have a small and statistically insignificant effect. Non-essential business closures and restaurant and bar limits are other policies that have a positive and statistically significant impact on presence at home, although their effect sizes are around half of what is observed for statewide stay-at-home orders.

**Table 2:** Effect of COVID-19 policies on community mobility

| VARIABLES | (1)<br>Presence at home<br>[0.32] | (2)<br>Grocery & pharmacy<br>[10.32] | (3)<br>Parks<br>[25.16] | (4)<br>Retail & recreation<br>[5.57] | (5)<br>Transit stations<br>[1.99] | (6)<br>Workplaces<br>[-0.73] |
| --- | --- | --- | --- | --- | --- | --- |
| Mean daily temperature (°F) | -0.079***<br>(0.009) | 0.120***<br>(0.030) | 1.596***<br>(0.143) | 0.154***<br>(0.030) | 0.149***<br>(0.029) | 0.074***<br>(0.019) |
| Statewide stay-at-home orders | 2.058***<br>(0.501) | -7.978***<br>(1.379) | -11.314**<br>(4.651) | -4.990***<br>(1.619) | -3.179*<br>(1.799) | -3.984***<br>(1.162) |
| Other stay-at-home orders | 0.515<br>(0.590) | -1.500<br>(2.311) | -3.567<br>(7.370) | -0.258<br>(2.267) | 0.493<br>(2.073) | -0.630<br>(1.121) |
| Non-essential business closures | 1.280***<br>(0.423) | -2.453**<br>(1.150) | 2.695<br>(4.490) | -3.146***<br>(1.088) | -4.876***<br>(1.755) | -4.710***<br>(0.976) |
| Large gathering bans | -0.064<br>(0.391) | 1.486*<br>(0.794) | 7.052<br>(5.869) | 1.875**<br>(0.906) | 4.073**<br>(1.838) | 0.959<br>(1.048) |
| School closure mandates | 0.339<br>(0.372) | -0.901<br>(1.151) | -4.565<br>(2.965) | -3.446**<br>(1.474) | -2.931**<br>(1.236) | -0.609<br>(0.948) |
| Restaurant/bar limits | 0.780**<br>(0.361) | -1.708<br>(1.048) | 1.749<br>(4.158) | -2.888***<br>(0.955) | -4.979***<br>(1.821) | -2.552***<br>(0.952) |
| Observations | 1,477 | 1,478 | 1,477 | 1,479 | 1,479 | 1,478 |
| R-squared | 0.977 | 0.939 | 0.674 | 0.982 | 0.957 | 0.978 |

Notes: Each column reports regression coefficients from a linear regression model, weighted by state population in 2018. In addition to the listed variables, we control for state and day-of-the-month fixed effects for each regression. Numbers in brackets are mean outcome variables before the implementation of the first social distancing policy. Negative means suggest there was a decline in those outcomes before the first social distancing policy. Standard errors in parentheses are clustered at the state level. \*\*\* $p < 0.01$ , \*\* $p < 0.05$ , \* $p < 0.1$

The rest of the table presents results on changes in visits from out-of-home places. Interestingly, they suggest a decline in all of these measures, providing evidence that the results obtained for presence at home are not spurious. Among all of the statistically significant estimated coefficients in Table 2, those related to statewide stay-at-home orders are the most pronounced ones, which strongly suggests that this particular policy is likely the most effective in promoting social distancing. On the other hand, policies such as large gathering bans seem to have a limited and statistically insignificant effect on keeping people at home. Table 3 provides estimates by adding state-specific day-of-week variables to each model. Overall, the results are very similar and suggest that state-specific day-of-week variations in outcomes are are not driving our results.

**Table 3:** Effect of COVID-19 policies on community mobility, results with state-specific day- of-week fixed effects

| VARIABLES | (1)<br>Presence at home<br>[0.32] | (2)<br>Grocery & pharmacy<br>[10.32] | (3)<br>Parks<br>[25.16] | (4)<br>Retail & recreation<br>[5.57] | (5)<br>Transit stations<br>[1.99] | (6)<br>Workplaces<br>[-0.73] |
| --- | --- | --- | --- | --- | --- | --- |
| Mean daily temperature (°F) | -0.075***<br>(0.009) | 0.125***<br>(0.030) | 1.536***<br>(0.150) | 0.148***<br>(0.030) | 0.156***<br>(0.032) | 0.073***<br>(0.020) |
| Statewide stay-at-home order | 2.015***<br>(0.532) | -7.624***<br>(1.336) | -11.742**<br>(4.968) | -4.768***<br>(1.636) | -3.092<br>(1.855) | -3.834***<br>(1.228) |
| Other stay-at-home orders | 0.532<br>(0.598) | -1.431<br>(2.272) | -4.478<br>(7.553) | -0.275<br>(2.204) | 0.387<br>(2.083) | -0.615<br>(1.130) |
| Non-essential business closure | 1.265***<br>(0.423) | -2.795**<br>(1.083) | 3.618<br>(4.498) | -3.355***<br>(1.070) | -4.865**<br>(1.823) | -4.813***<br>(0.968) |
| Large gatherings ban | -0.024<br>(0.400) | 1.387*<br>(0.788) | 6.605<br>(6.079) | 1.875**<br>(0.895) | 3.992**<br>(1.880) | 0.935<br>(1.070) |
| School closure mandate | 0.314<br>(0.392) | -0.828<br>(1.165) | -4.068<br>(3.108) | -3.348**<br>(1.521) | -3.042**<br>(1.277) | -0.564<br>(1.002) |
| Restaurant/bar limits | 0.795**<br>(0.365) | -1.756<br>(1.093) | 2.220<br>(4.363) | -2.971***<br>(0.975) | -4.927**<br>(1.873) | -2.697***<br>(0.983) |
| Observations | 1,477 | 1,478 | 1,477 | 1,479 | 1,479 | 1,478 |
| R-squared | 0.978 | 0.944 | 0.687 | 0.983 | 0.958 | 0.979 |

Notes: Each column reports regression coefficients from a linear regression model, weighted by state population in 2018. In addition to the listed variables, we control for state and day-of-the-month fixed effects, and state-specific day-of-week fixed effects for each regression. Numbers in brackets are mean outcome variables before the implementation of the first social distancing policy. Negative means suggest there was a decline in those outcomes before the first social distancing policy. Standard errors in parentheses are clustered at the state level. \*\*\* $p < 0.01$ , \*\* $p < 0.05$ , \* $p < 0.1$

Next, we provide evidence for the dynamic effects of the policies of interest on presence at home, in Figure 3. Overall, except for *other stay-at-home orders* and to some extent, *non-essential business closures*, there are no differences in presence at home trends before policies took effect between the states with and without those policies. This is evident through the flat trends before the policies took effect on day zero. Consistent with the results in Table 2, we observe the largest effect on presence at home through statewide stay-at-home orders, although the magnitude of the effect declines after a week. The effects of other stay-at-home orders and non-essential business closures are noisy. Large gathering bans illustrate flat trends before and after the implementation, suggesting that this policy is ineffective in changing individuals’ behavior in terms of staying at home. Finally, both school closures and restaurant and bar limits seem to positively affect presence at home. However, their effects are either weak and marginally significant or there is some evidence of upward trends before the policy implementation.

**Figure 3:**
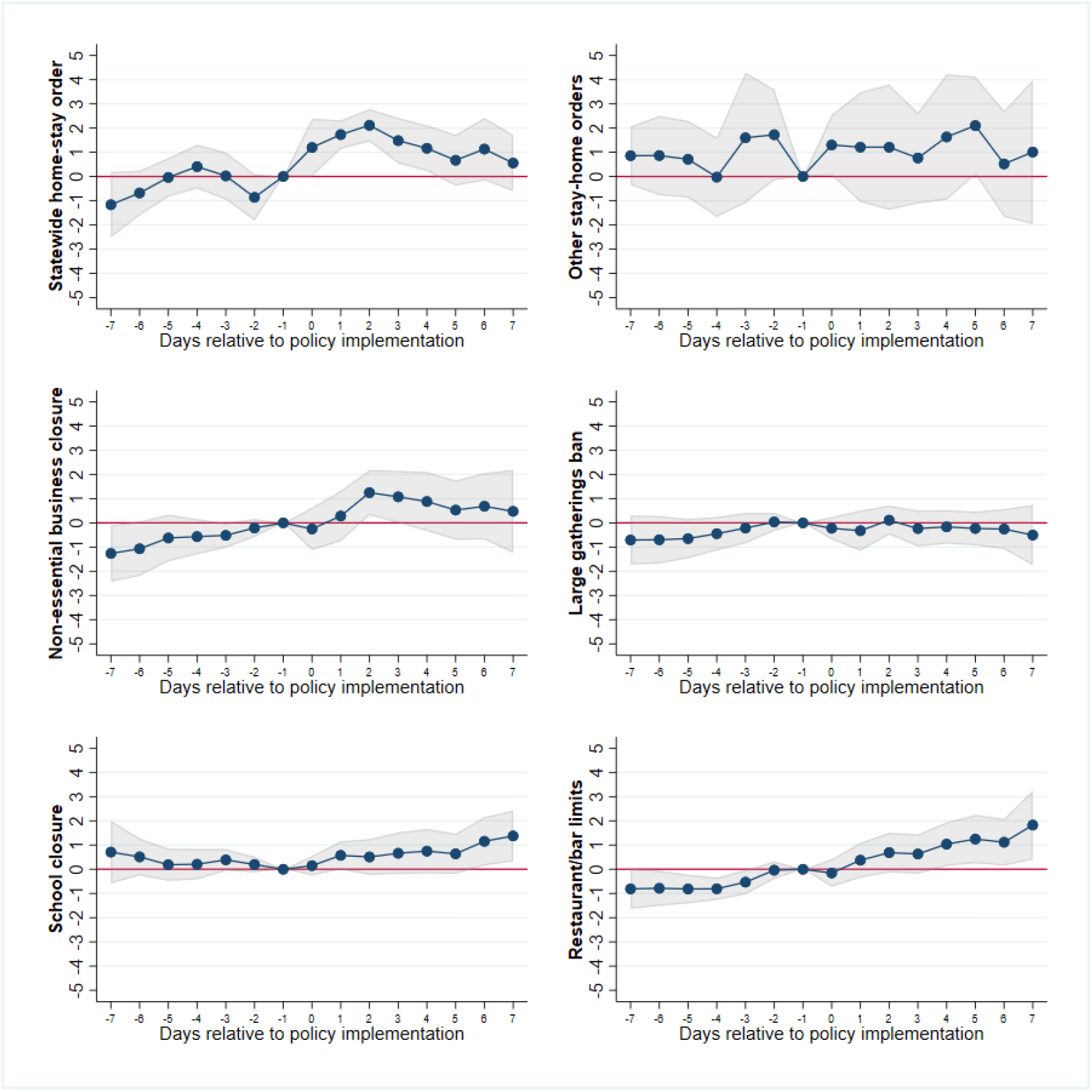
Event study of policies of interest on presence at home. Gray areas highlight the 95% confidence intervals.

The preexisting trends in the outcome in states that adopt other stay-at-home orders and non-essential business closures provide further evidence supporting the role of voluntary actions (possibly driven by public awareness factors before any policy implementation) in changing stay-at-home behavior. It is worth noting that we considered the start date of each policy as the first day in which the policy was in effect for 24 hours. This assumption can impact what we see on the last day before the policy in the event study graphs. Therefore, changes immediately before the policy date must be interpreted carefully.

Given that COVID-19 started to spread in the US from the states of Washington, California, and New York, and that these states experienced a higher volume of positive cases and deaths, there is always a concern that the estimated policy effects are driven by these states. We provide a version of a permutation test in which we drop each state from the sample, one at a time, then estimate the effect of the statewide stay-at-home order. The estimated coefficients were consistent when dropping each state, suggesting the effects are not driven by a particular group of states. Results presented in Figure 4 support this hypothesis.

**Figure 4:**
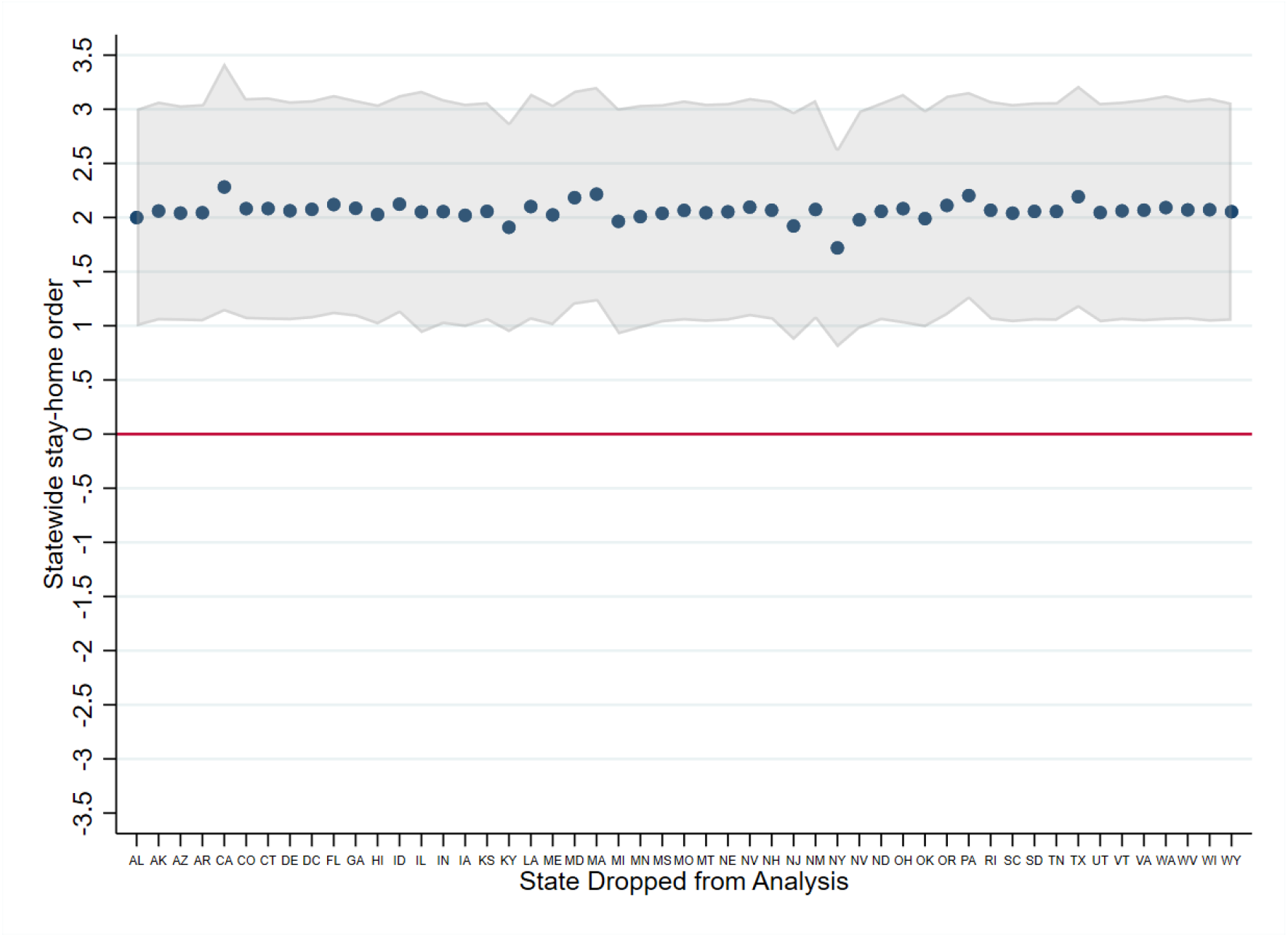
Sensitivity of estimates to dropping states one at a time, community mobility.

### Impact of Policies on Daily Positive Cases

We now turn to test the impact of social distancing policies on the dynamic of the disease based on publicly reported positive test results (confirmed cases). Our approach is similar to what we did for mobility analysis, but here we use a Poisson regression model in an event-study framework.

Based on our mobility results, we expect that the stay-at-home policy results in a significant reduction in the number of confirmed cases after a delay that accounts for the period from infection to test results. This is confirmed by our event-study analysis (Figure 5), where we see a steady decline in the number of daily confirmed cases starting from 10 days after the policy date that gets to a 37% reduction in the number of daily cases after 15 days, compared to the baseline scenario with no such policy. Note that a minimum of 10-day delay is consistent with several reports that estimate the median incubation period of the new Coronavirus in a range between 4 to 5 days (Lauer et al., 2020; Guan et al., 2020) and the way testing has so far been handled in the US where most tests (85% as of the second week of April 2020) are conducted at private laboratories with several days of wait time for the result (“Why some covid-19 tests in the US take more than a week”, 2020), and with current CDC testing guidelines (“Testing in the U.S”, 2020) that recommend prioritizing hospitalized patients, which further prolongs the delay between infection and test results.

**Figure 5:**
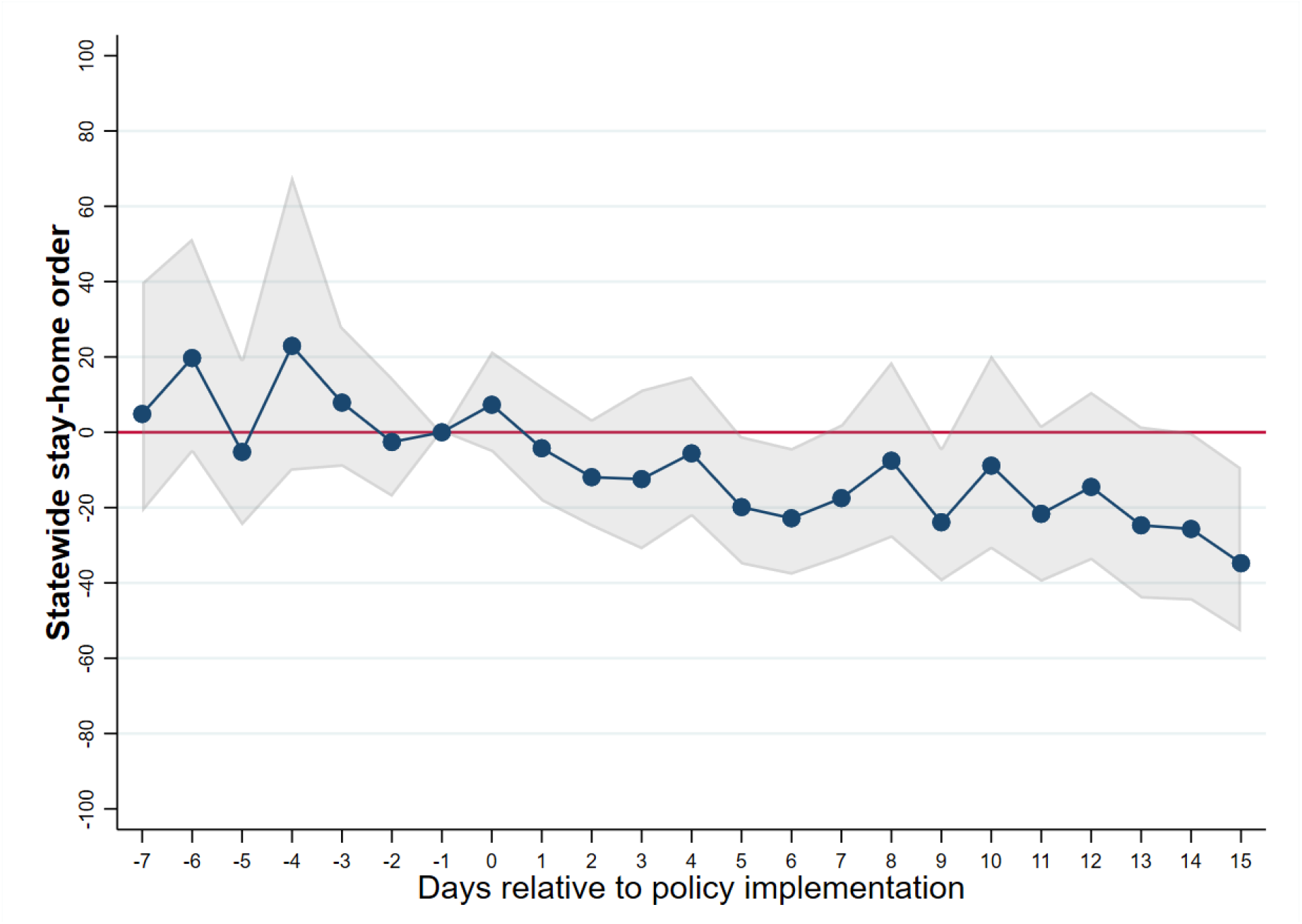
Event study of statewide stay-at-home order on positive test frequencies. Gray areas highlight the 95% confidence intervals.

Similar to the analysis we conducted for community mobility, we drop each state one at a time and estimate the effect of the statewide stay-at-home order. For this analysis, each estimated coefficient captures the effect of the policy at least 15 days after the policy implementation. This specification is more inline with findings in event study analysis. Results reported in Figure 6 suggest that the effect of this policy on confirmed cases is consistent and does not depend on inclusion of specific states in the sample.

**Figure 6:**
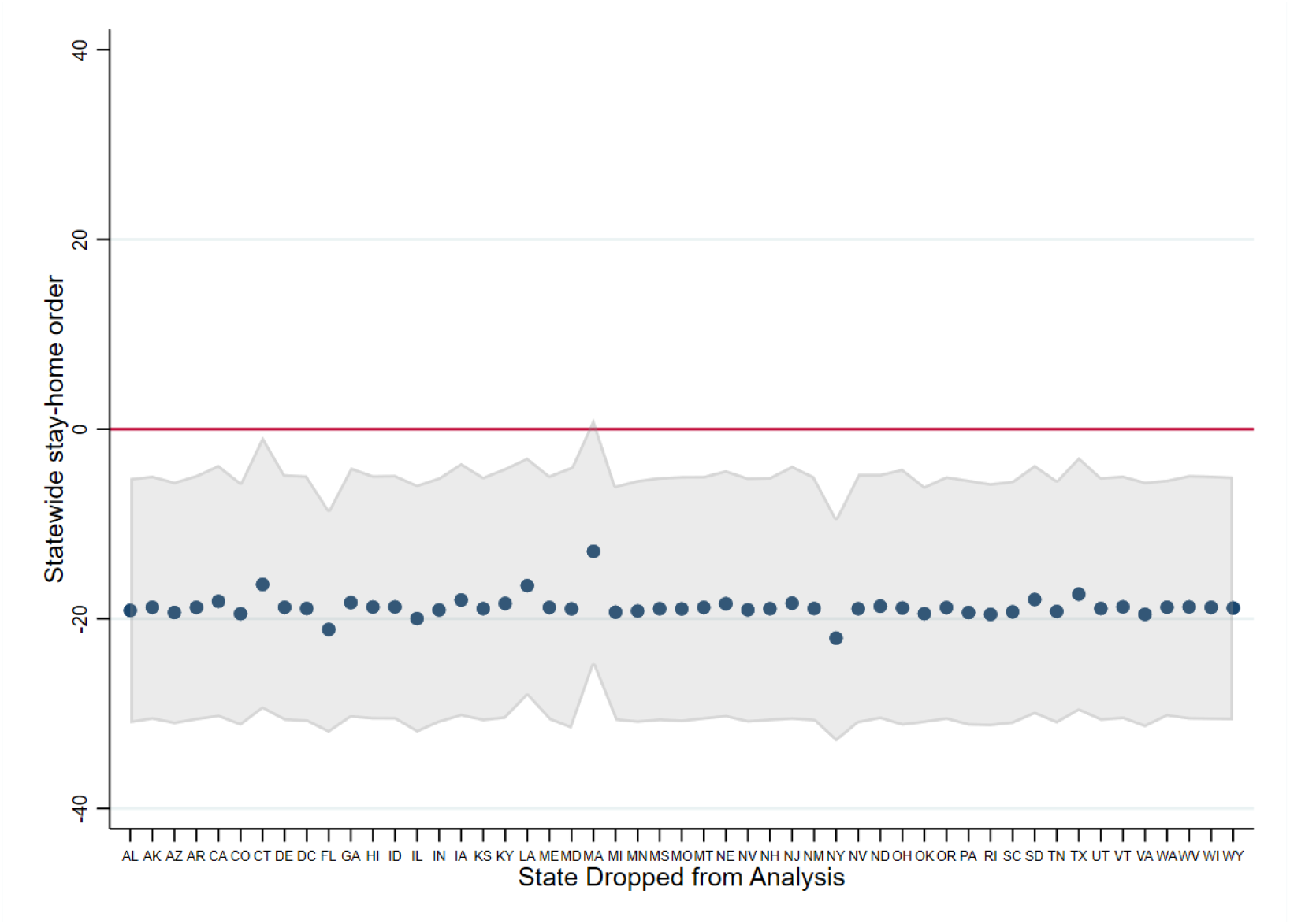
Sensitivity of estimates to dropping states one at a time, confirmed COVID-19 cases.

Figure 7 presents the event study results for other COVID-19 policies. Overall, as expected from the mobility-based analysis in the previous section, there are relatively small and statistically insignificant effects after the implementation of those policies.

**Figure 7:**
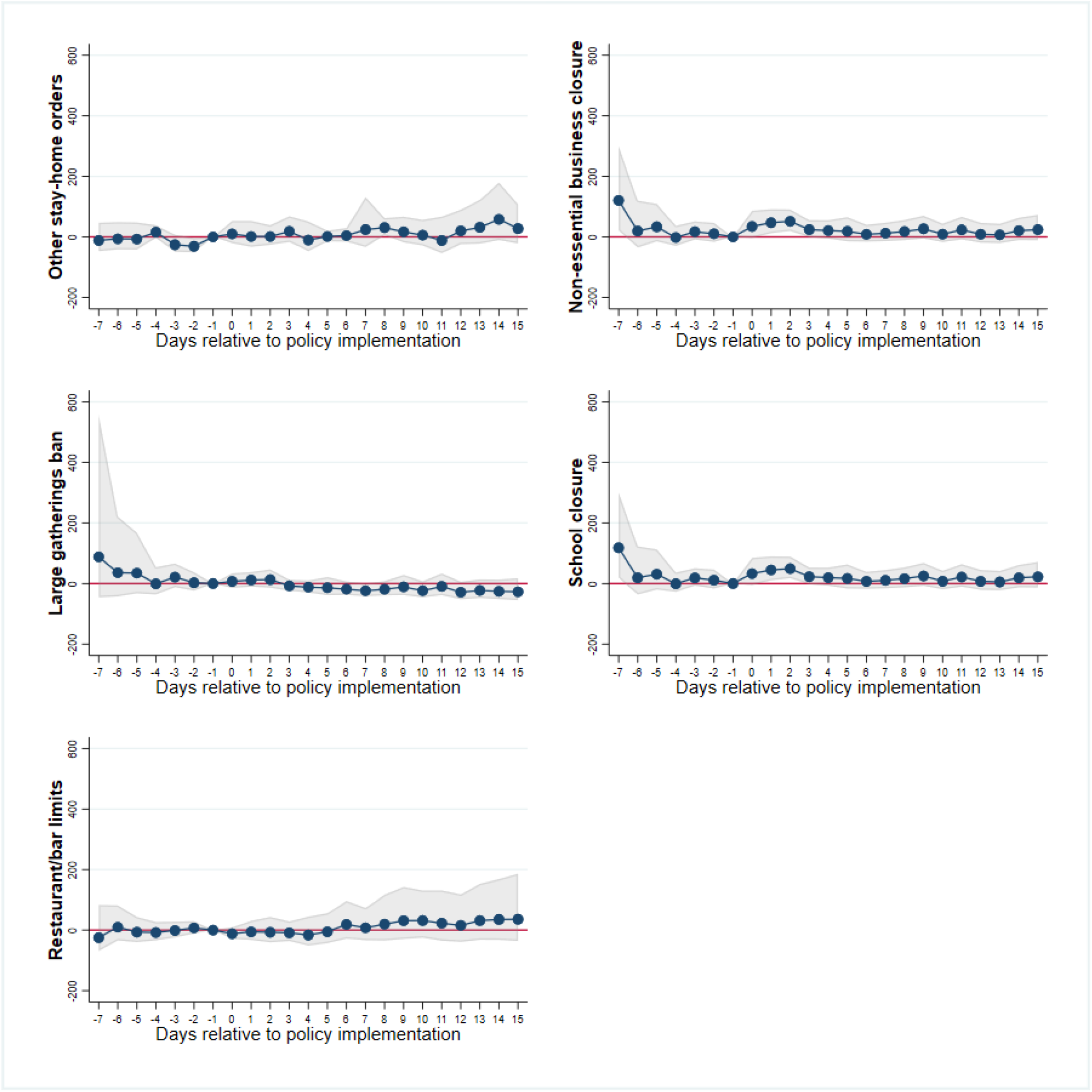
Event study of COVID-19 policies on positive test results. Gray area highlights the 95% confidence intervals.

## Discussion and Conclusion

Our findings show the effectiveness of different social distancing policies on reducing out-of-home social interaction during the early stage of the COVID-19 outbreak. We show that reductions in out-of-home social interactions are driven by a combination of policy and voluntary measures, then demonstrate the strong causal impact of state-wide stay-at-home orders and the more moderate impact of non-essential business closures and bar/restaurant limits. At this stage of the outbreak in the US, other policy measures such as school closure mandates or large gathering bans seem to have had no significant causal impact on keeping people at home. We need to be cautious when generalizing the results from this early stage of the pandemic to the later stages and to possible future waves of the outbreak. Specifically, we need to emphasize that our results do not claim that more lenient social distancing policies such as school closures or large gathering bans are always causally inefficient in reducing social interaction. While it is evident that most of the social distancing *capacity* of such measures is already absorbed in non-policy driven changes–possibly caused by social awareness–it is expected that as the pandemic lasts longer, voluntary social distancing measures will start to wane, making such policies (individually or in combination) more effective in later stages of the pandemic (N. Ferguson et al., 2020).

Our study has a number of limitations. First, the Google database is not based on the universe of all smartphone users and it only includes those individuals who have enabled the Location History setting on their account. However, given that around 90 percent of users keep their location services on (“Smartphone Users Keep Location Services Open”, 2020), our estimates should not be largely affected. Similarly, the data are imperfect since they don’t include people without smartphones and those who don’t carry their phones to certain places. However, this should not affect *changes* in recorded behavior and is expected to have little impact on our results.

Finally, it is worth noting that measuring the effectiveness of social distancing policies based on confirmed cases hinges on how the tests are conducted in different states, requiring more than controlling for just the number of conducted tests that we used in this study. This problem can be in part mitigated by analyzing the policy effects on the number of COVID-19 deaths. Given that the median time from infection to death is reported to be close to 17 days, and since many states have issued their strongest policies in the last week of March, that study needs to wait until enough reliable data are collected.

## Data Availability

This article uses publicly available data with sources listed under the references.

## Footnotes

1 Here we assume that large indoor gatherings at residential places have not increased substantially since the start of the outbreak.

